# Dolodoc, a mobile application for the self-management of chronic pain: Acceptability and usability study

**DOI:** 10.1101/2025.10.28.25338623

**Authors:** Julie Guebey, Laetitia Gosetto, Benno Rehberg, Christian Lovis, Frederic Ehrler, Aude Molinard-Chenu

## Abstract

**Background:** Approximately 19% of adults in Europe are affected by chronic pain, which reduces quality of life. Pain management apps (mHealth) offer a promising solution for self-management, but users engagement and adherence can be a limitation to their clinical impact. User experience design and studies play an important role in optimizing usability and long-term adoption of digital health interventions.

**Objective:** This study aims to evaluate the user experience of Dolodoc, a mobile application for chronic pain self-management, using a mixed-methods approach that evaluates acceptability through a content quality survey and examines usability by analyzing overall usage patterns.

**Methods:** A cross-sectional acceptability study was conducted among chronic pain patients recruited from the Geneva University Hospitals pain center and through snowball sampling. Participants rated 84 evidence-based self-management strategies using a five-point Likert scale based on 5 acceptability criterias: understandability, motivation, feasibility, relevance, and alignment with the related quality-of-life dimension. Usability was assessed through usage metrics that were collected over six months using Piwik PRO analytics to observe the usage behaviors of real-world Dolodoc users.

**Results:** In the acceptability study, a total of 33 participants rated the self-management strategies positively across all dimensions. On a scale from −2 to 2, the strategies were well understood (mean = 1,47), motivational (1.12), feasible(1.01), relevant (0.99), and aligned with the dimensions (1.33). The usability study demonstrated that 60% of patients used Dolodoc only once, indicating that long-term adherence remains a challenge. Within Dolodoc, pain tracking, useful links and medication logging were the most actively used features.

**Discussion:** This study highlights the gap between acceptability and long-term adherence to mHealth solutions. Improving personalization and accessibility could increase user engagement and long-term adherence. Future iterations of the app should incorporate tailored interventions and real-time feedback mechanisms. In addition, taking advantage of a digital navigation follow-up could facilitate user adoption and sustained engagement.

## INTRODUCTION

Chronic pain is a major health care problem that affects about 19% of adults in Europe^1^. This pernicious symptom impacts patients globally and on many dimensions of their quality of life^2^, underscoring the need to develop new approaches to care. Indeed, chronic pain often forces individuals to explore multiple approaches to alleviate the pain^3^. Managing chronic pain involves an integrative medicine approach based on pharmaceutical or non-pharmaceutical treatments, psychological support and self-management strategies^4^. Optimizing self-management is paramount in the promotion of self-efficacy^5^, which stands out as a powerful lever^6^ to improve clinical outcomes related to chronic pain such as disability, affective distress and pain severity^7^.

In this context, mobile digital health (mHealth) solutions represent a promising sector^8^ and there is a growing number of smartphone applications that are developed to take advantage of these innovations in the care trajectories of patients suffering from pain^9,10^. In fact, the benefits of mobile applications for chronic pain management have been demonstrated, showing effects on quality of life and pain severity^11,12^. However, we postulate that their effectiveness depends on how well they address the needs and perceptions of the target users. Indeed, motivational aspects are key in the engagement of patients toward self-management^13^ and multimodal treatment^14^. Yet, long-term engagement and adherence have emerged as key limiting factors for digital interventions in medicine^15,16^. One way to increase the adherence and engagement of target users is User experience (UX) design, it aims to clarify the understanding of the human-product interaction, identifying key components and dimensions to improve design practices^17^. It represents an essential framework at every stage of the design process, aiming to enhance the overall quality of the final product, based on the needs of target users^18^. UX design in health innovations falls within the contemporary concept of person-centered medicine which prioritizes patient engagement and individual care ^19,20^. Commonly used methods in UX studies include usage metrics analysis, user interviews, usability testing, surveys, and behavioral analytics. These methods can be applied at various stages of the innovation process, ensuring adaptability to different research and design needs. Mixed methods studies facilitate the understanding of UX, allowing for better adoption and increased effectiveness of digital mHealth tools^21^. Globally, UX studies aim to understand two main characteristics of a mHealth solution: acceptability and usability ^22^.

Dolodoc is a mobile application that was designed to facilitate self-management of chronic pain. It features pain self-management strategies focusing on seven quality-of-life dimensions, the possibility to add information about the user’s pain, and to share a summary of pain-related activity and outcomes with healthcare professionals^23^. A recent study shows the significant gaps between the functionality being developed and end-user expectations, highlighting the need for close collaboration between designers, developers and users throughout the co-design process ^24^.

In the present study, we are describing the perception of users about the content of Dolodoc through five acceptability criteria that were selected, based on previous literature: understandability^25^, motivation^26^, feasibility^26^, relevance^27^, and alignment with the related quality-of-life dimension. Usability will be assessed by observing the usage habits of real-world users. Overall, this work aims to describe important UX outcomes of a mHealth solution for chronic pain through a mixed method approach.

## METHODS

### Study design

This mixed method cross-sectional study assesses UX outcomes about a chronic pain self-management mobile application, including acceptability through content quality assessments by users and usability through global usage metrics. This study was approved by the local ethical committee for human research.

### Mobile application description

Dolodoc is a mobile application that was developed to assist patients in the self-management of chronic pain. It provides self-management strategies and encourages users to maintain a digital journal to dynamically monitor their pain and related quality-of-life indicators. The overall development and implementation of this digital tool has been described elsewhere^23^ and has been following the UX design criteria for optimization. The core content of the app consists of a library of 84 evidence-based self-management strategies that users can test in real life, tracking their effects within the app on various pain severity indicators, including quality-of-life measures. The strategies were developed through consultations with a focus group of chronic pain patients and professionals specializing in chronic pain care. They first discussed the impact of pain and how it affected their daily life in different aspects. From these discussions, seven quality-of-life dimensions emerged: mood, social support, sleep, work, intimacy, relaxation and daily activities. Drawing on these dimensions and scientific evidence, a team of psychologists developed motivational pain-management strategies: each quality-of-life dimension refers to 4 to 20 strategies that are encompassed in a corpus of 84 self-management strategies. This content was proofread by a pain specialist and by a communication expert.

### Participants

Participants of the content evaluation survey were eligible for inclusion if they were 18 or older, suffered from chronic pain, were fluent in french, and were able to provide informed consent. Participants were mainly recruited from the pain center of the University Hospital of Geneva, but other patients meeting inclusion criteria were also recruited through snowball sampling.

There was no requirement to download or use the application to participate, allowing the study to include feedback from experienced users, novices, and non-users alike. To ensure that each participant fully understood the framework within which the strategies were provided, a contextual presentation of the mobile application was given before participants shared their opinions.

### Setting, recruitment and data collection

#### Acceptability study: content quality assessment by users

Recruitment took place from June to September 2024. Some participants were recruited passively through flyers made available in the waiting room, but most of them were recruited actively by members of the pain center team. All 84 strategies were randomly divided into five groups, in order to reduce the total duration of the survey : based on the time of their recruitment, participants were allocated to a random sample of 5 to 22 strategies.

Participants were asked to fill out an online questionnaire including demographics, pain characteristics (duration and impact on the seven quality-of-life dimensions that are included in Dolodoc) and an evaluation of each sampled strategy. Each strategy was assessed by 5 to 10 participants according to five acceptability criteria: understandability, motivational impact, feasibility, relevance, and alignment with the related quality-of-life dimension. Responses were collected on a five-point Likert scale ranging from “strongly disagree” to “strongly agree”, allowing for a quantitative understanding of participants’ perceptions of each strategy.

### Usability study: real-world usage metrics

Usage data were obtained from Piwik PRO analytics Suite (Piwik PRO, 2024), an online usage analytics platform that collects Dolodoc usage metrics from anonymous data tracking with cookies and sessions data. These data reflect the behavior of all Dolodoc users during the whole study period, not just about the participants who participated in the evaluation interviews. Data were collected over a 6-months period, from January to June 2024.

A cookie (named _pk_id) is collected to determine if a user is recurring and to calculate the number of visits and the duration of each session. Another cookie (named _pk_ses), is used to indicate whether a session is active. The information contained in these cookies is sent to Piwik at each event during the usage of Dolodoc (e.g. a page view).

The unique identifier from _pk_id cookies was used as a proxy for identifying unique visitors, while session data were extracted from _pk_ses, including operating system, session number, title of the visited page, time spent on the page, session duration. Although most of our analyses include both iOS and Android users, the number of sessions per visitor could not be calculated for iOS users, likely because on iOS _pk_id is deleted after 30 minutes of inactivity due to Apple’s stricter cookie management. As a result, Piwik always identifies iOS users as unique new visitors, preventing us from distinguishing recurring visitors from unique visitors.

### Statistical methods

Descriptive statistics and inferential statistics were applied depending on the nature of data. The following software was used for data analyses and visualization: Microsoft Excel, Jupyter Notebook^28^, Orange Data Mining^29^.

## RESULTS

### Acceptability study

#### Demographics and pain-related characteristics

A total of 33 participants (20 females, 13 males) were enrolled in the study. 3 participants evaluated 2 different subsamples of strategies; hence the total number of assessments is up to 36 per question in the survey. For these three recurrent participants, we minimized intra-individual variability by using only their most recent response in demographics variables.

The participants’ ages ranged from 23 to 79 years old, with a mean age of 38.1 ± 15.63 years.

Most participants reported chronic pain for more than one year (24/33), while 9 participants reported chronic pain for less than one year.

Each quality-of-life dimension was affected in various intensities, but overall, quality-of-life dimensions were not homogeneously affected by chronic pain in our sample (Khi^2^ = 21,4, ddl = 24, p = 0.00025) (Figure 1). These results support the idea of an individual profile in the impact of pain on quality of life.

**Figure 1:**
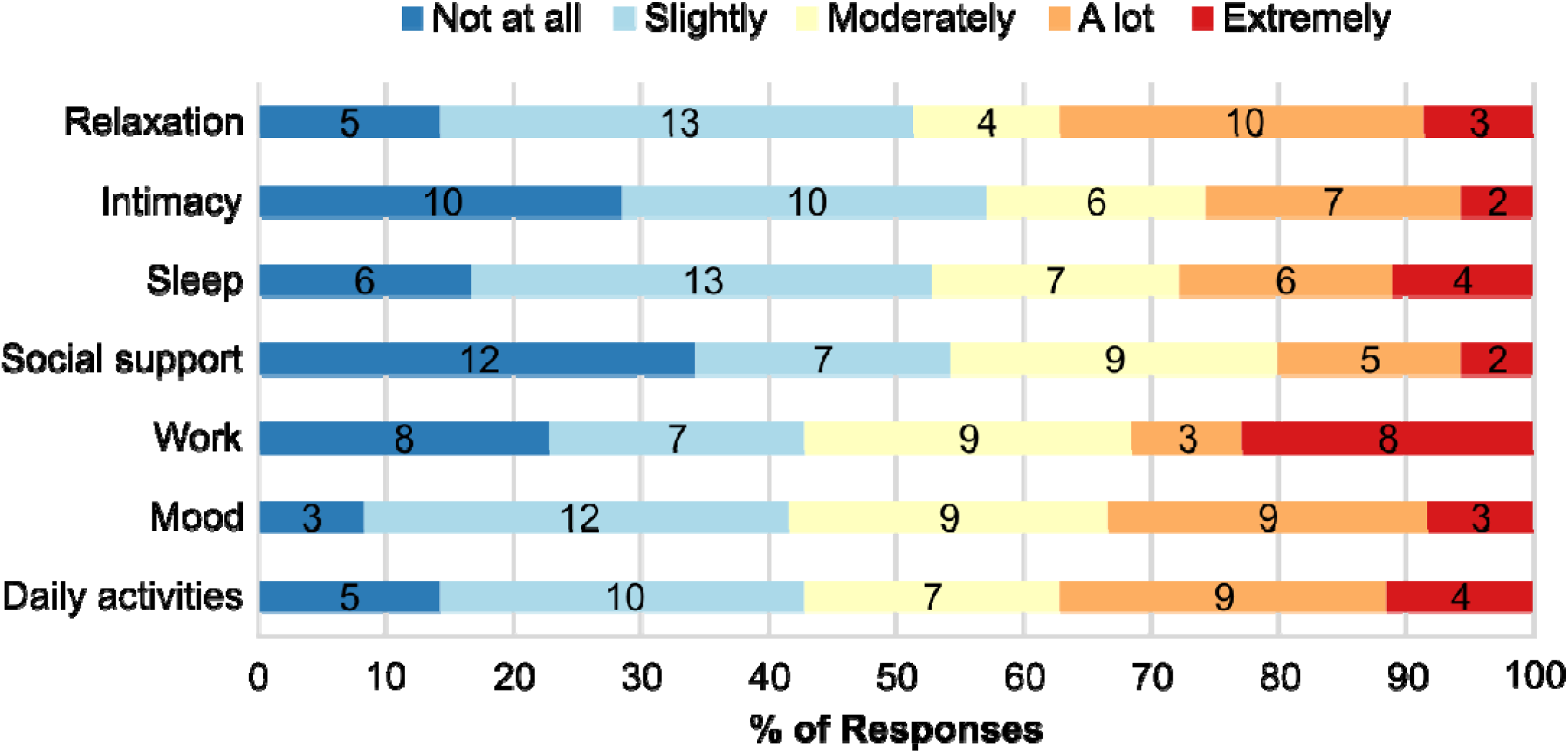
Impact of pain on the seven dimensions of quality of life that are targeted in the self-management strategies incorporated in Dolodoc mobile app, as reported by participants. n = 33 participants, the number of assessments in each subfraction (35-36 in total) is indicated within the bars, 5 missing values were excluded.

#### Content evaluation survey

Overall, the content of Dolodoc was well perceived by participants: on a Likert scale from −2 to 2 (from strongly disagree to strongly agree) the average rating on all acceptability criteria was 1.18. Moreover, 62/84 strategies received a strong agreement (mean > 1), whereas 22/84 were agreed upon (mean > 0), demonstrating that acceptability criteria are globally fulfilled according to our user sample. More specifically, understandability was the highest-rated acceptability criterion (mean = 1.47) while relevance (mean = 0.99) and feasibility (mean = 1.01) received the lowest ratings. The strategies were also perceived as motivational (mean = 1.12) and aligned with the specific dimensions (mean = 1.33) (Figure 2.A). Each acceptability criterion received between 610 and 617 ratings. In this study, missing values represented 0.27% of data, because the setting of the online questionnaire did not require to reply to all questions before submitting the survey.

**Figure 2:**
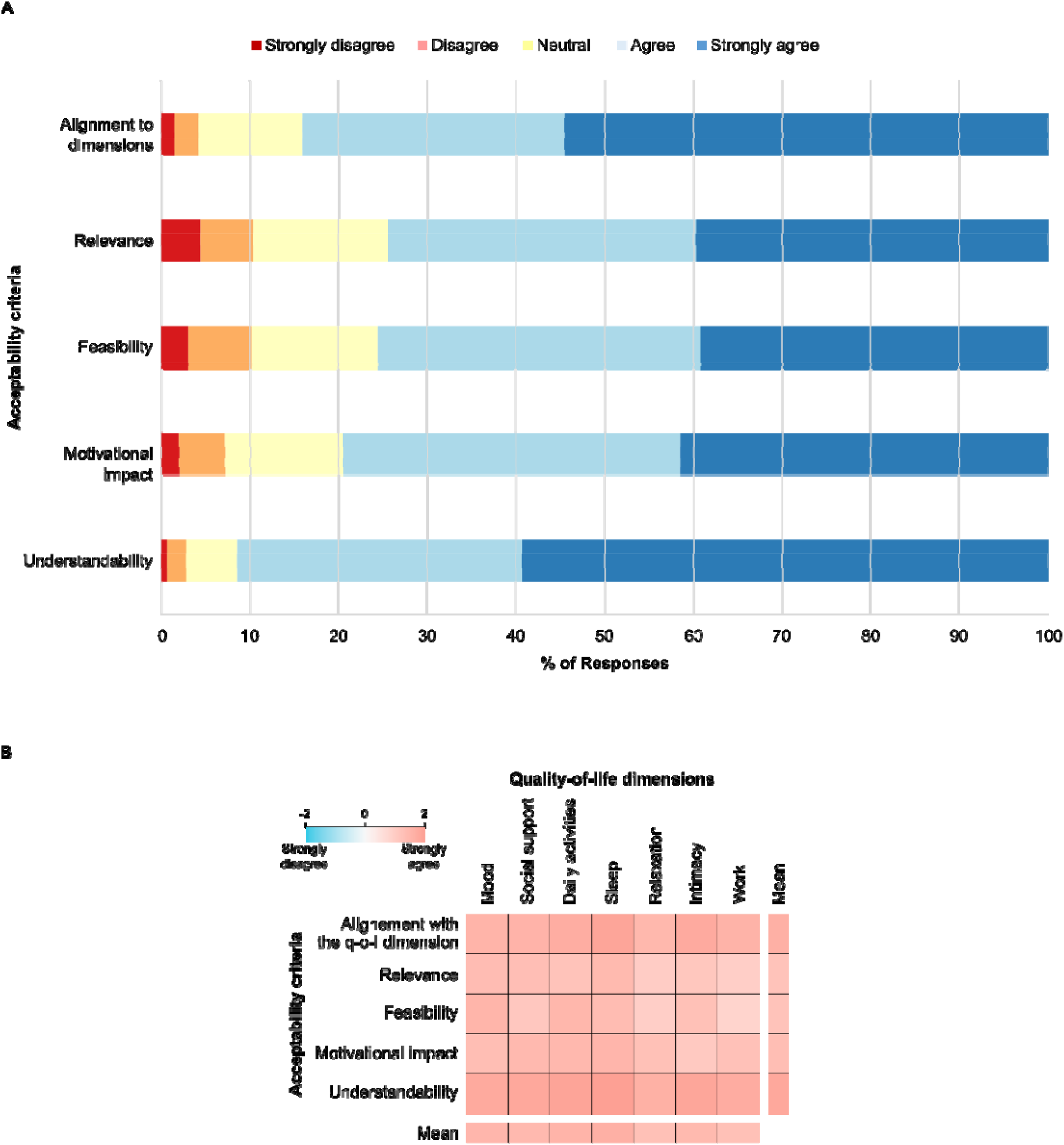
Pain management strategies in Dolodoc are well perceived by users across five acceptability criteria and seven quality-of-life (q-o-l) dimensions. A. Bar plot showing the distribution of ratings across five acceptability criteria. B. Heat map displaying the average rating score for acceptability criteria and q-o-l dimensions. n = 3077 ratings in total.

To precisely assess the perception of the strategies in the context of well-being, we calculated a mean rating for each of the seven previously described quality-of-life dimensions, based on each acceptability criterion (Figure 2.B). The mean scores range between 1 (agree) and 2 (strongly agree), ranging from 1.08 for the ‘relaxation’ dimension and 1.40 for the ‘sleep’ dimension. The detailed numbered results are provided in supplementary table 1. These results suggest a positive consensus among participants regarding Dolodoc self-management strategies within all quality-of-life dimensions.

### Usability study

Over a 6-months period, from January to June 2024, a total of 3,589 unique sessions were recorded, with 59% using iOS and 41% Android operating systems. The mean duration of each session was 3 minutes and 21 seconds (201 seconds) (Figure 3.A). In average, 10.39 page views occurred per session. We spotted 0.07% of missing values in the page titles data, because these events were not considered as a page view. Missing values were excluded from analysis. The bounce rate (i.e. the percentage of sessions where only one page view occurred) was 23.29 %.

**Figure 3:**
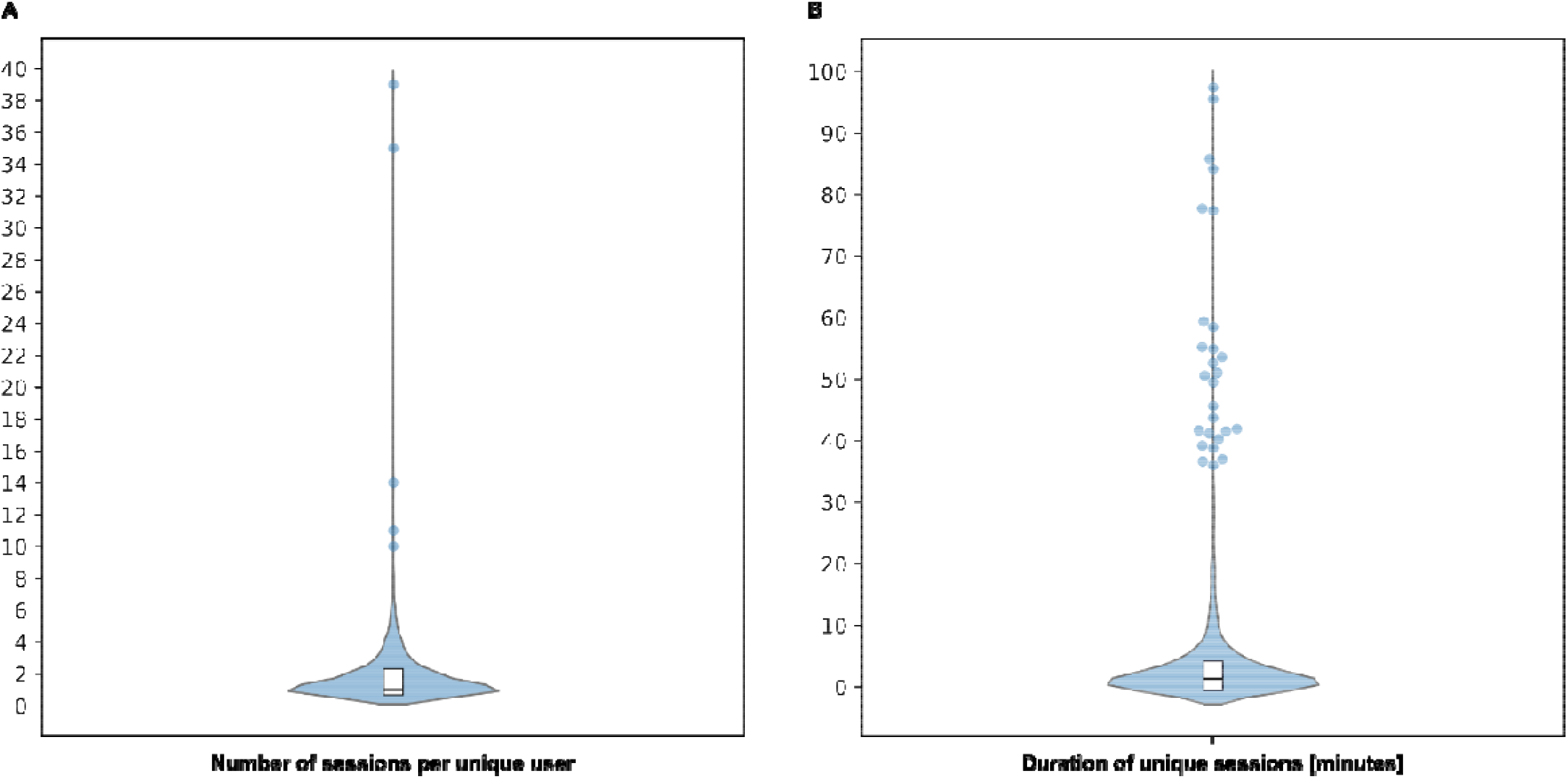
Analysis of user interaction with Dolodoc based on sessions number and duration. Violin plots showing the distribution of the number of sessions per unique user (A) and the duration of each unique session (B). Box plots show the interquartile range and the median. (A) n = 1473 sessions from Android users; (B) n = 3,589 sessions from Android and iOS users. Outliers (above 95^th^ percentile) are displayed as individual points.

Among Android users, a total of 1,473 sessions were observed for 802 unique visitors. 60.6% of these visitors visited the application only once and the mean number of sessions per visitor was 1.84, although a few outliers display more than 10 sessions over this period (Figure 3.B). As mentioned in the methods section, iOS data were not available for this analysis.

Table 1 displays the top 10 most visited pages by all users. When navigating through the application, the most visited page was the “first launch page”, showing again the significant proportion of users who mostly used Dolodoc once. Next, visitors most often viewed the home page and the page where they can monitor quality-of-life dimensions through a 4-points visual scale. Interestingly, the page where the strategies are compiled is the first action in the mobile app that users perform actively, because the 3 previous pages are automatically displayed at connection. These results are coherent with the architecture and the purpose of Dolodoc, leading users towards self-observation and exploring the included strategies for the self-management of chronic pain.

**Table 1:** Top 10 most visited pages in Dolodoc over 6 months. Q-o-l = Quality of life.

| Page | Number of views | Fraction of total views [%] | Functionality | Type of user interaction |
| --- | --- | --- | --- | --- |
| First launch | 9830 | 26.40 | Welcome | Passive |
| Home | 5737 | 15.41 | Welcome and Monitoring | Passive |
| Q-o-l dimension evaluation | 4333 | 11.64 | Monitoring | Passive |
| Strategies | 4312 | 11.58 | Management | Active |
| Report export | 2860 | 7.68 | Monitoring | Active |
| Details of activities | 2509 | 6.74 | Management | Active |
| User profile | 1479 | 3.97 | Personalization | Active |
| Graphical view of collected data | 843 | 2.26 | Monitoring | Active |
| History of pain | 789 | 2.12 | Monitoring | Active |
| Dashboard | 661 | 1.78 | Monitoring | Active |

Interestingly, the time spent on each page of the application does not follow the same architectural logic as the most visited pages (Table 2) : users spent most of the time on less accessible pages of the application like a page where they can describe and add characteristics to their pain named “add your pain”, a page containing useful links, and a page where they can add their current medication named “add medication”. These results show what users actively seek when they navigate through the application, providing precious feedback and understanding of their expectations and needs.

**Table 2:** Top 10 most engaging pages in Dolodoc based on average visit duration over six months.

| Page | Average time spent [seconds] | Application content functionality | Type of user interaction |
| --- | --- | --- | --- |
| Add your pain | 78.97 | Personalization | Active |
| Useful links | 59.70 | Education | Active |
| Add medication | 48.15 | Personalization | Active |
| Association | 39.65 | Social support | Active |
| Home | 39.08 | Welcome and Monitoring | Passive |
| Graphical view of collected data | 28.89 | Monitoring | Active |
| Create an activity | 28.64 | Management | Active |
| Report | 23.71 | Monitoring | Active |
| Print report | 21.31 | Monitoring | Active |
| Personal informations | 20.32 | Monitoring | Active |

## DISCUSSION

### Key results

This study provides a detailed evaluation of user experience in the context of a pain-management mobile application (Dolodoc), focusing on acceptability, through quality assessment of its content by a sample of users, and usability through an analysis of user interactions with the application.

The population that evaluated the content of the application was unevenly affected by pain, when assessing its impact on seven quality-of-life dimensions, highlighting the variability in the experience of chronic pain and the need for personalized approaches in pain management.

Self-management strategies that are included in Dolodoc were perceived as engaging by users, based on five acceptability criteria: understandability, motivational aspect, feasibility, relevance and alignment to dimensions. This result was not straightforward, because even though the application was developed using a user-centered design, end-users were not involved in the redaction of this content^23^. Real-world usage monitoring of Dolodoc over 6 months demonstrated that 60% of Dolodoc users visit the application once and that long-term adherence remained a challenge. However, users actively engaged with less accessible features like personal pain and medication description, underscoring the importance of being active using the application, but also the need of personalization, to enhance engagement and motivation^27^. Pages containing educational and social support were also among the most actively visited pages within the application, highlighting their relevance. Overall, this mixed-method study provides a balanced UX report on a mobile application for the self-management of chronic pain.

### Limitations

The main limitation of the acceptability study is the relatively small number of patients (N=33). Additionally, patients were recruited mostly by healthcare professionals, which can lead to a selection bias with the most motivated patients participating. However, our sample was diversified, in terms of demographics and pain-related characteristics. Another limitation of the acceptability study is the assessment of tailored acceptability criteria, instead of a standard measurement instrument such as the uMARS scale^32^. However, our study focuses on a french-speaking population and at the time of the conception of the study the uMARS scale was not translated into french. As mentioned in the methods sections, the main limitation in the usability study was the exclusion of iOS users from the number of sessions per visitor calculation.

### Interpretation

This study highlights the challenges of adopting a pain self-management application, identifying an important gap between acceptability and long-term engagement. This rare combination of real-world and experimental UX observations in the context of chronic pain provides useful patient-centered insights that could help improving Dolodoc and inspire similar mHealth innovations. Indeed, post-production UX studies in the context of mHealth solutions for chronic pain are sparse and are usually based on a small sample of users, lacking real-world usage data^24,31,34,35^. Using both ecological and survey-based data helps mitigating the “trial bias” that has been previously described as an important confounder in user engagement studies in the study of unguided e-health interventions^33^.

Users are satisfied about the content of this mHealth solution, suggesting that the user-centered design that guided its development^23^ lead to a good acceptability of Dolodoc, similarly to other mHealth interventions for chronic pain. In comparison, other therapeutic modalities, such as CBT^38^, mindfulness-based interventions^39^, physical activity^40^ and multimodal group therapy^41^ are also generally well accepted by patients suffering from chronic pain. However, real-world usage habits in our study demonstrate the need to further enhance long-term engagement. Poor long-term engagement has already been described for other mHealth interventions in chronic disorders^16^ and chronic pain^42^, highlighting important barriers for use such as the lack of reminders^26,30^.

The diversity of pain profiles among our sample of users in the acceptability study, as well as the fact that global users actively spend most of their time on pages that are dedicated to personalization features reveal the importance of personalizing pain management mHealth solutions, in accordance to previous reports^25,26^. The personalization of mobile health applications positively influences users’ trust, thereby increasing the likelihood of their adoption^43^.

### Generalizability

Despite the aforementioned limitations, the study provides a strong initial assessment of acceptability and usability of Dolodoc. Although these UX results are specific to this mHealth solution, our results are in line with other reports about similar interventions in terms of usability^44,45^ and acceptability^37^.

### Perspectives

Based on the results of the present study, it seems relevant to add features aiming at improving engagement over time, such as a reward system and more personalization features^26^. Additionally, it would be useful to revise the app navigation logic to facilitate access the most frequently used pages. Future improvements could also help us collect UX data in a longitudinal manner, for example by adding the possibility for users to rate features within the app, by “liking” our “disliking” the content of the application.

However, optimizing digital therapeutic alliance^46^ remains a key challenge for the future of mHealth. In this regard, digital navigation follow-ups have emerged as a potential solution, based on human interactions, to maximize the engagement of patients toward mHealth solutions^47^ and increase their clinical impact^48^.

## Data Availability

All data produced in the present study are available upon reasonable request to the authors

## Acknowledgements

We would like to thank Myriam Perrier for her help navigating the usability study results

## Authors contributions

JG collected data, analyzed data and contributed to the writing of this publication (data visualizations, original draft, revising and editing). AMC supervised the project and contributed to the writing of this publication (data visualizations, original draft, revising and editing). FE supervised the project and contributed to the writing of this publication (revising and editing). LG contributed to the writing of this publication (original draft, revising and editing) and supervised JG. CL supervised LG, JG and AMC and contributed to the writing of this publication (revising and editing). BR helped recruiting patients and contributed to the writing of this publication (revising and editing).

## Notes

### Competing Interest Statement

The authors have declared no competing interest.

### Funding Statement

The Private Foundation of the HUG of the Geneva University Hospitals funded the developpement of the application.

### Author Declarations

Business Administration System for Ethics Committees of Cantonal Research Ethics Committee waived ethical approval for this work.

